# Children with Autism using the Floreo Virtual Reality Building Social Connections Module: a Feasibility Study

**DOI:** 10.1101/2025.08.28.25334679

**Authors:** Kevin Shapiro, Vijay Ravindran, Gregory Downing, Shirley Mak-Parisi, Georgia Barbayannis, Sinan Turnacioglu

## Abstract

Immersive virtual reality (VR) technology shows promise as a therapeutic aid to support social communication in children with autism spectrum disorder (ASD). The main objective was to assess the feasibility of using Floreo’s VR headset with the BSC module curriculum as a component of behavioral therapy for preschool and school-age autistic children. Using a randomized control study design, a total of 14 participants (8 participants in the intervention group and 6 participants in the control group) received approximately 36 sessions of Floreo’s BSC treatment or VR control up to 3 times a week over a 12-15 week period in an in-clinic ABA therapy setting. Outcomes were measured using the validated and reliable Autism Impact Measure (AIM), which provides a composite score as well as subdomain scores for communication, social reciprocity, peer interaction, repetitive behavior, and atypical behavior. As a primary outcome measure, we evaluated change from baseline in AIM composite and subdomain scores as a function of treatment. The Floreo VR headset was well-tolerated by study participants and was incorporated without difficulty into clinical treatment sessions. No serious adverse events occurred, and no participants dropped out of the study due to undesirable side effects. Autistic children who received Floreo’s immersive VR BSC program showed an overall mean improvement in AIM composite score (-25) compared to those in the control group (-0.84) at a clinically meaningful level, although this did not achieve statistical significance. There was a clinically and statistically significant improvement in the AIM communication score for children in the Floreo BSC group (-5.12) compared to the control group (+3.33, p=0.02). The study findings suggest that Floreo’s Building Social Connections Module is safe and well-tolerated, and has the potential to enhance social communication skills and reduce challenging behaviors in autistic children.

## Introduction

### Background on ASD

Autism spectrum disorder (ASD) is a pervasive, heterogeneous neurodevelopmental condition defined by impairments in communication and social skills, as well as repetitive and restricted behaviors and interests.^1^ While the diagnosis is often made in childhood, the impact of autism is lifelong, including reduced social engagement and quality of life.^2^ Caregivers of autistic individuals experience increased levels of strain, which has been exacerbated by the COVID-19 pandemic.^3,4^ The prevalence of ASD is increasing; it is estimated that in 2020, 1 in 36 eight-year-olds in the United States met criteria for the diagnosis.^5^ In this manuscript, “autism,” “autism spectrum disorder,” “on the autism spectrum,” “autistic individual,” and “individual with autism” will be used interchangeably as they are the terms preferred by community groups.^6,7^ While much attention has been paid to the pathogenesis and timely diagnosis of ASD, the rapidly increasing prevalence of the disorder has not been matched by an increase in the availability of therapies. Evidence-based early interventions for individuals with ASD, including applied behavior analysis (ABA) and naturalistic developmental behavioral interventions,^8-10^ have been shown to produce significant financial and social cost savings for families and for society as a whole.^11^ However, access to early behavioral intervention services varies markedly depending on geographic, socioeconomic, and other demographic factors.^12-15^ Even when it is available, children with autism can show significant variability in their response to early intervention.^16,17^

As a result of these discrepancies in access and response to existing therapies, many children diagnosed with autism continue to experience serious social, cognitive, language, and other difficulties as adults.^18,19^ These difficulties negatively impact quality of life, social and community integration, and emotional and affective functioning for both autistic individuals and their family members. Families of young adults who were initially diagnosed with ASD as preschoolers have noted the persistence of unmet social needs^20^ and increased caregiver stress.^21^ A survey of young adults with ASD found that almost one-third reported social isolation,^22^ and other studies have observed high rates of anxiety and depression.^23,24^ Roux et al.^25^ reported that 4 in 10 young adults with ASD were disconnected from school and work through their late teens and early twenties, and less than 20% of young adults with ASD lived independently. Some studies have suggested that as autistic children age into adulthood, they may experience worsening deficits compared to peers in social communication^26^ and adaptive functioning.^27^

The increasing population prevalence of autism, coupled with limitations in the availability and efficacy of existing evidence-based therapeutic approaches, has created a pressing need for effective, low-cost interventions that address core symptoms of autism in children.^28^ Developing effective interventions for social communication in particular is critical for public health, given the high financial and psychological costs of social disengagement for autistic individuals and their families. In addition, supporting developmental skill training is among the highest priorities for research identified by the autism community, including clinicians, individuals with autism, and family members.^29,7^

### Background on VR

Immersive Virtual Reality (VR) has the potential to fill a critical treatment gap for individuals with ASD and related disorders, particularly beyond the early intervention period. Researchers have already begun to explore the potential of VR technology for targeting ASD-related deficits, ^30^ and VR approaches have proven effective therapeutically for children with other psychiatric challenges, including specific phobias and panic disorders.^31-33^

To date, most therapy-focused VR research in autism has not been immersive.^34-36^ In contrast to a non-immersive experience, immersive VR gives the user the feeling of being inside a virtual world, and offers an environment in which users can try out experiences that are hard to stage or replicate in real life. Modern immersive VR incorporates visual, spatial, and auditory elements that are motivating for individuals with and without ASD,^37^ and include programmable contingent reward animations that can be “gamified.”

Immersive VR technology is emerging as a highly effective therapeutic tool for school-aged children with ASD,^38,39^ offering them a safe environment in which to develop independence. Several studies have begun to explore the benefits of immersive VR therapy for ameliorating symptoms of ASD,^40-42^ but limitations in study design have made the results difficult to generalize to a community setting. The present study was developed to evaluate the effectiveness of an immersive VR intervention for social communication using a design that addresses some of the limitations of prior studies. In particular, we aimed to conduct the study in a naturalistic setting in which all participants were receiving standard-of-care therapy (ABA) for autism as a baseline. We also sought to employ more clinically relevant outcome measures that can be analyzed longitudinally.

### Information about Floreo

Floreo’s VR platform integrates wireless tethered VR screens (iPhones) and tablets (iPads) that are seamlessly linked in real-time, allowing the therapist (or monitor) to set up lessons, change the interactive environment, and provide live verbal and VR-based guidance, feedback, and rewards to users. The platform uniquely allows parents, caregivers, and educators to observe and engage with the child’s learning experience, enhancing the overall therapeutic process.

Floreo’s immersive VR experience incorporates automated activities, actions, and rewards, which support efficient and effective VR scene navigation, interaction, and learning. Studies have shown improvements in emotional regulation in children with ASD with immersive virtual reality.^43^ After each lesson, a results card is provided to the monitor based on the performance from the user. This approach enhances traditional therapist-mediated interventions by incorporating immersive VR environments to boost user engagement and learning.

In a previous pilot study focused on joint attention using Floreo’s VR system with school-age autistic students, the joint attention module proved to be feasible, safe, and well-tolerated, with no participants reporting side effects or dropping out due to undesirable side effects.^39^ Moreover, improvements were seen in joint attention-related skills, specifically an increase in total number of interactions, enhanced use of eye contact, and more frequent initiation of interactions.^39^

## Materials and Methods

In this study, immersive VR was used to train social skills in preschool- and school-aged children with ASD. The primary objective was to test the feasibility of this intervention in a naturalistic clinical setting. To this end, we monitored the safety and tolerability of Floreo BSC over the course of a 12-15 week treatment period. We also evaluated the efficacy of Floreo’s Building Social Connections (BSC) module (henceforth Floreo BSC) for ameliorating core symptoms of autism, namely, deficits in social communication and social interaction, assessed by parent report using the validated Autism Impact Measure (AIM). AIM scores were compared between children receiving Floreo BSC and those in a control group who were also exposed to VR but did not participate in the BSC curriculum.

### Protection of Human Subjects

The study was reviewed and approved by WCG, the Central Internal Review Board.

### Participants

Participants were recruited from among children receiving ABA therapy at five clinical sites operated by Cortica, a network of clinical centers that provide multidisciplinary services for children with neurodevelopmental disabilities such as ASD. In addition to ABA, Cortica centers provide developmental therapies, including speech-language therapy and occupational therapy, as well as neurologic and neurobehavioral medical care. Conducting the study at Cortica sites therefore ensured to the extent possible that all participants were already receiving comparable standard-of-care interventions for autism. Specific inclusion criteria and exclusion criteria are listed in Table 1.

**Table 1.** Inclusion and Exclusion Criteria for Study Participants.

| Inclusion Criteria | Exclusion Criteria |
| --- | --- |
| Children aged between 4-10 years old with a documented diagnosis of ASD<sup>a</sup> based on the clinical judgment of a qualified clinician according to DSM-5<sup>b</sup> criteria, supported by either the Autism Diagnostic Observation Schedule or the Autism Diagnostic Interview-Revised | History of photosensitive epilepsy or known photosensitive response on electroencephalogram |
| Participants received ABA<sup>c</sup> therapy at Cortica | Active diagnosis of migraine headache or migraine variant (abdominal migraine, cyclic vomiting syndrome) not controlled by medication or other therapy |
| Tolerability of the VR<sup>d</sup> therapy | Vertigo, motion sensitivity, ataxia, or other serious balance disorder |
| - | Primary sensory impairment such as blindness or deafness |
| - | Motor disorder that would interfere with VR<sup>d</sup> engagement |

<sup>a</sup>ASD, autism spectrum disorder
<sup>b</sup>DSM-5, Diagnostic and Statistical Manual of Mental Disorders, Fifth Edition
<sup>c</sup>ABA, Applied Behavior Analysis
<sup>d</sup>VR, virtual reality

### Study Design

This study was designed as a randomized controlled trial of an intervention to be administered in the context of in-clinic ABA therapy. Participants were randomized 1:1 to an active treatment or VR control group. Parents or caregivers of participants, assessors, and study investigators were blinded to assignment to participant and assignment to treatment or control arms. Participants and study staff administering the treatment were not blinded, as there were non-trivial experiential differences between the treatment and control conditions.

Participants who met inclusion and exclusion criteria (Table 1) participated in 36 sessions of treatment or VR control 3 times a week over a 12-week period, with up to 3 additional weeks to make up for any missed sessions. The sessions lasted approximately 15 minutes and took place in a clinic setting with the assistance of a member of the study staff acting as a VR monitor. The VR monitor was a trained ABA specialist who also assisted with headset adjustment, program activation, and monitoring.

#### Procedure

This study consisted of three phases: (i) screening and randomization; (ii) treatment; and (iii) follow-up (Figure 1).

**FIG. 1.** Schematic of the study design.

##### Screening visit

During the screening visit, conducted either in person at a clinic or virtually via Zoom, informed consent was obtained from a parent or caregiver of each potential participant, and informed assent was sought from potential participants before initiating any study-specific procedures.

Demographic and clinical characteristics of participants were collected, including age, biological sex, race, co-occurring diagnoses, medications, and concomitant therapies. The study coordinator asked the child to wear the VR headset to check for tolerability. If the child was able to wear the headset, a VR video lasting 4-5 minutes was played, and the study coordinator completed a post-session questionnaire to determine if the child could tolerate the VR intervention. A participant was judged to tolerate the intervention if they were able to complete the video without becoming agitated or removing the headset.

##### Baseline visit

At the baseline visit, the Autism Impact Measure (AIM) clinical scale was administered by qualified trained raters. While this paper focuses on the AIM, other clinical scales were also administered, including the Vineland Scales of Adaptive Behavior, 3rd Edition (Vineland-3), Childhood Autism Rating Scale, 2nd Edition (CARS-2), and Kaufman Brief Intelligence Test, 2nd Edition (KBIT).

##### Randomization phase

Following the baseline visit, participants were randomized into either treatment or control arms. The randomization of subjects to study groups was performed using a randomization scheme reviewed and approved by an independent statistician.

As noted above, parents and caregivers were blinded as to whether participants received Floreo VR therapy versus the active VR exposure control. Participants and study staff administering the sessions were not explicitly told whether they were assigned to Floreo VR therapy or control exposure, but there were unavoidably clear experiential differences between the two conditions. Research staff assisting with session logistics were not blinded. However, the clinicians completing pre- and post-intervention assessments, most of the sponsor team, and the statistician who performed the analysis were all blinded to group assignment.

##### Treatment phase

During treatment visits, participants received either treatment with the Floreo Building Social Connections (BSC) module or VR control under the supervision of a member of the study staff who acted as a VR coach (Figures 2 and 3). All VR coaches were registered behavior technicians (RBTs) who had experience working with autistic children in a clinical setting.

**FIG. 2.** An example of the monitor’s view of a lesson from Floreo’s Building Social Connections Module.

**FIG. 3.** An example of the monitor’s view of a lesson from Floreo’s Building Social Connections Module.

Table 3 depicts the lesson skills categories and lesson plans. Sessions (including VR lessons and questions) lasted approximately 15 minutes. Following each session, the VR coach completed a post-session questionnaire with the participant regarding the tolerability of the session, whether the session was completed, and whether any technical issues occurred.

**Table 2.** MCT^a^ Thresholds for Clinical Improvement and Deterioration for Each AIM^b^ Domain.

| **AIM<sup>b</sup> Domain** | **Clinical Improvement**<br>(95% CI<sup>c</sup>) | **Deterioration**<br>(95% CI<sup>c</sup>) |
| --- | --- | --- |
| AIM<sup>b</sup> Total | -4.49<br>(-7.61, -1.37) | 9.86<br>(5.12, 14.59) |
| Social Reciprocity | -0.68<br>(-1.25, -0.10) | 1.09<br>(0.21, 1.97) |
| Communication | -0.89<br>(-1.15, -0.28) | 1.53<br>(0.59, 2.46) |
| Peer Interaction | -0.89<br>(-1.44, -0.34) | 1.61<br>(0.76, 2.45) |
| Repetitive Behavior | -0.10<br>(-0.96, 0.77) | 1.47<br>(0.15, 2.79) |
| Atypical Behavior | -1.09<br>(-1.82, -0.37) | 1.76<br>(0.66, 2.86) |

<sup>a</sup>MCT, meaningful change threshold<sup>b</sup>AIM, Autism Impact Measure<sup>c</sup>CI, confidence interval

**Table 3.** Lesson Skills Category for Feasibility Curriculum.

| **Skills Category** | **Number of Lessons** |
| --- | --- |
| Communicative Gestures | 12 |
| Language Comprehension | 6 |
| Motor Imitation/Body Awareness | 9 |
| Nonverbal - Communicative Eye Gaze | 17 |
| Social Communication/Interactions | 28 |

The intervention included two lessons from the Floreo BSC program, each lasting 4-5 minutes. These VR lessons were delivered through the Floreo app, installed on an iPhone worn by the participant in a VR headset. The VR experience was managed and operated by the VR coach, who controlled the interaction via a linked iPad and was physically present with the participant during the session.

Control sessions consisted of two non-interactive VR videos, each lasting 4-5 minutes. VR episodes were presented via YouTube videos played on an iPhone worn by the participant in the VR headset. As with the Floreo BSC intervention, the control VR interaction was managed and operated by the VR coach, who was physically present with the participant in the clinic.

Participants randomized into the active VR exposure control engaged in sessions at the same frequency as the Floreo BSC arm. The study included a total of 36 planned treatment visits over 12 weeks, with visits scheduled to occur 3 times per week. An extension in duration of the study up to 15 weeks was allowed to make up for missed sessions due to illness, holidays, and family obligations. Participation in the study ended after 15 weeks even if participants had not completed all 36 treatment phase visits.

##### Interim visits

Study staff conducted interim visits with the parent or caregiver of each participant after treatment sessions 12 (interim visit 1), 18 (interim visit 2), and 24 (interim visit 3). At interim visits 1 and 3, parents or caregivers were asked about any adverse events that may have occurred since the screening visit. At interim visit 2, the AIM was completed.

##### End-of-study visit

At the end of the study, the parent or caregiver was asked about any adverse events that may have occurred since the last interim visit, and the AIM was completed.

### Assessments

#### Autism Impact Measure (AIM)

The primary outcome measure in this study was the AIM, a reliable and validated 41-item parent questionnaire targeting sensitivity to changes in core ASD symptoms.^44,45^ The AIM was specifically designed to assess treatment outcomes and symptom improvements in children with ASD over a short interval.^46^ The questionnaire uses a 2-week recall period with items rated on two corresponding 5-point scales (frequency and impact). The items cover distinct, empirically-derived subdomains of ASD symptoms: namely, repetitive behavior, atypical behavior, social reciprocity, communication, and peer interaction.^46^ A composite score is also calculated.^46^ The subdomains and total scores have been shown to be sensitive both to overall changes in a child’s condition and to changes resulting from different treatment conditions.^46^

The clinical significance of changes in AIM scores can be interpreted at the within-person level using the meaningful change threshold (MCT), which relates absolute changes in scores to real-world improvement or worsening in symptoms.^47^ MCT values for improvement and deterioration shown in Table 2 were estimated using anchor-based, caregiver-reported perceptions of change in a large-scale study of caregivers of 2,761 autistic children, aged 3-17 years old, over a 12-month period.^47^

The AIM was completed online by the parent or caregiver of each participant and took approximately 30 minutes to complete.

### Measures of safety

Safety was measured by recording all adverse events reported by study staff or parents or caregivers of the participants. Adverse events could include changes in health or behavior related to the study as well as exacerbations of known medical or behavioral conditions, effects of concomitant medications, or other changes reported by parents, caregivers, or study staff. Serious adverse events were defined as those that were life-threatening, required hospitalization, or resulted in death or disability.

### Data Analysis

The primary endpoint for this study was change in composite score on the AIM. Secondary endpoints included change in individual AIM subdomain scores. All outcomes were presented using descriptive statistics to evaluate the distribution of key variables and relevant covariates between treatment and control groups. To evaluate the success of stratified randomization procedures, Fisher’s exact test for categorical variables was performed to evaluate differences in the variables included in stratification.

The primary analysis compared change from baseline to post-test in the AIM composite score across study groups (Floreo VR vs VR control) in order to evaluate the effectiveness of Floreo VR Building Social Connections Intervention on autism symptoms. This was performed using a multilevel mixed model with participants included as a random effect and AIM composite score included as the dependent variable. A cross-level group by time interaction was included as the primary coefficient of interest. Secondary analyses evaluated change over time for each of the AIM subdomain scores (social reciprocity, communication, peer interaction, repetitive behavior, and atypical behavior). To evaluate the effectiveness of the intervention, *post hoc* pairwise comparisons compared group-level scores at end-of-study.

## Results

A total of 31 participants gave informed consent/assent and were screened for this study. One participant in the control group completed some study visits but withdrew from the study early as the caregiver decided that they no longer wished to participate; no adverse event was reported.

A total of 14 participants were included in the final analysis, comprising 6 participants in the control group and 8 participants in the Floreo BSC treatment group (Table 4). All enrolled participants were retained in the intent-to-treat (ITT) analysis if they completed an end-of-study visit, regardless of the number of interim visits completed.

**Table 4.** Descriptive Study and Characteristics of Study Participants.

|  | **Control** | **Treatment** |  |
| --- | --- | --- | --- |
|  | (*n* = 6) | (*n* = 8) | ***p*-Value** |
|  | Mean<br>(SD<sup>a</sup>)/(%) | Mean (SD<sup>a</sup>)/(%) |  |
| Age (months) | 68 (14) | 72 (21) | 0.9 |
| Gender |  |  | >0.9 |
| Female | 1 (17%) | 1 (13%) |  |
| Male | 5 (83%) | 7 (88%) |  |
| Race (%) |  |  | >0.9 |
| Asian | 2 (33%) | 4 (50%) |  |
| Other | 1 (17%) | 1 (13%) |  |
| White | 3 (50%) | 3 (38%) |  |
| Ethnicity |  |  | >0.9 |
| Hispanic or Latino | 3 (50%) | 3 (38%) |  |
| Not Hispanic or<br>Latino | 3 (50%) | 5 (63%) |  |
| Vineland-3 Group (%) |  |  | 0.2 |
| Mild-Moderate | 3 (50%) | 7 (88%) |  |
| Severe | 3 (50%) | 1 (13%) |  |

<sup>a</sup>SD, standard deviation

The mean age of participants was 68 ± 5.7 months in the control group, and 72 ± 7.4 months in the treatment group (Table 4). Consistent with broader ASD demographics, the majority of participants were males, with one female in the control group and one female in the treatment group. In terms of the severity of autism, 50% of the control group participants and 88% of the intervention group participants had mild to moderate autism based on Vineland-3 scores (Table 4).^48,49^ There were no significant group differences in race, ethnicity, or autism severity between groups (Table 4).

We also examined the exposure of participants to ABA and developmental therapies over the course of the study. All participants in both groups received concomitant ABA and speech therapy. There were non-significant differences in the proportion of participants in each group receiving occupational therapy (OT), speech therapy (ST), physical therapy (PT), and music therapy (MT). The average frequency of weekly therapy sessions did not vary significantly except for music therapy, which skewed towards higher usage in the treatment group (Table 5).

**Table 5.** Current Therapies and Co-morbidities.

|  | **Control**<br>(*n* = 6) | **Treatment**<br>(*n* = 8) | ***p*-Value** |
| --- | --- | --- | --- |
| ABA<sup>a</sup> | 6 (100%) | 8 (100%) |  |
| OT<sup>b</sup> | 6 (100%) | 6 (75%) | 0.5 |
| ST<sup>c</sup> | 6 (100%) | 8 (100%) |  |
| PT<sup>d</sup> | 4 (67%) | 1 (13%) | 0.091 |
| MT<sup>e</sup> | 5 (83%) | 5 (63%) | 0.6 |
| ABA<sup>a</sup> Frequency<br>(sessions/week) | 4.67 (1.03) | 3.63 (0.69) | 0.073 |
| OT<sup>b</sup> Frequency<br>(sessions/week) | 2.00 (0.63) | 2.00 (0.63) | >0.9 |
| ST<sup>c</sup> Frequency<br>(sessions/week) | 2.17 (0.41) | 1.94 (1.08) | 0.3 |
| PT<sup>d</sup> Frequency<br>(sessions/week) | 1.13 (0.25) | 1.00 (NA) | >0.9 |
| MT<sup>e</sup> Frequency<br>(sessions/week) | 1.10 (0.22) | 1.80 (0.45) | 0.038 |

<sup>a</sup>ABA, applied behavior analysis<sup>b</sup>OT, occupational therapy<sup>c</sup>ST, speech therapy<sup>d</sup>PT, physical therapy<sup>e</sup>MT, music therapy

During the study, three adverse events were reported in the control group, all involving the same participant whose results were included in the analysis. The first was an ear infection reported at interim visit 1. The second and third events were reported at the end of study visit and included a laceration requiring stitches and an asthma exacerbation. None of these events were judged to be related to the intervention. This participant dropped out of the study.

Two adverse events were reported in the treatment group. The first event occurred for a participant during the interim visit and was described as “increased mood swings” possibly related to the study intervention. The second event occurred for a different participant whose caregiver reported “increased yelling behavior” at the end of study visit, possibly related to the study intervention. These participants remained in the study and were included in the final analysis.

No serious adverse events were reported in either the treatment or the control group.

### Autism Impact Measure

There were no significant baseline differences in the AIM composite or subdomain scores at baseline between control and treatment groups (Table 6).

**Table 6.** Baseline Descriptive Statistics and Group Differences.

|  | **Control**<br>(*n* = 6) | **Treatment**<br>(*n* = 8) | ***p*-Value** |
| --- | --- | --- | --- |
|  | Mean (SD<sup>a</sup>)<br>[min-max] | Mean (SD<sup>a</sup>)<br>[min-max] |  |
| AIM<sup>b</sup> |  |  |  |
| Composite | 218.67 (56.72) [133.00-282.00] | 192.88 (48.85) [133.00-253.00] | 0.3 |
| Social | 26.00 (6.32)<br>[20.00-34.00] | 23.00 (5.90)<br>[15.00-32.00] | 0.3 |
| Communication | 35.17 (11.09)<br>[22.00-51.00] | 30.00 (7.13)<br>[22.00-40.00] | 0.4 |
| Peer Interaction | 25.00 (5.55)<br>[15.00-32.00] | 18.13 (6.71)<br>[10.00-27.00] | 0.08 |
| Repetitive Behavior | 38.17 (14.77)<br>[19.00-56.00] | 38.00 (11.64)<br>[19.00-52.00] | >0.9 |
| Atypical Behavior | 32.00 (11.22)<br>[16.00-46.00] | 27.13 (10.40)<br>[15.00-41.00] | 0.3 |

\*Indicates a significant test statistic at $p < 0.05$
<sup>a</sup>SD, standard deviation
<sup>b</sup>ABA, Applied Behavior Analysis

The primary outcome measure for this study was change in AIM composite score between treatment and control groups. On average, the AIM composite score improved by -25 points in the treatment group (surpassing the MCT for overall improvement), and by 0.84 points in the control group (no change with respect to MCT). Despite this numerical difference, the primary outcome analysis showed that there was no significant interaction between time and treatment group on the AIM composite score (Table 7). This indicates that the difference between groups in change from baseline to end-of-study was not statistically significant.

**Table 7.** Primary Outcome Measure Analysis.

|  | Control<br>(n = 6) |  | Control<br>Group<br>Change<br>Score | Treatment<br>(n = 8) |  | Treatment<br>Group<br>Change<br>Score | Interaction F-Statistics |
| --- | --- | --- | --- | --- | --- | --- | --- |
|  | Pre-Mean<br>(SD<sup>a</sup>) | Post-Mean<br>(SD<sup>a</sup>) |  | Pre-Mean<br>(SD<sup>a</sup>) | Post-Mean<br>(SD<sup>a</sup>) |  |  |
| AIM<sup>b</sup> Composite | 218.67<br>(56.72) | 217.83<br>(57.67) | -0.84 | 192.88<br>(48.85) | 167.88<br>(35.99) | **-25** | 1.50<br>(p=0.25) |

\*Indicates a significant Group×Time (pre-post) interaction test statistic at $p < .05$
†Indicates a significant estimated marginal means for post-hoc pairwise comparison test of within-group change at $p < 0.05$
**Bold** indicates an average decrease in AIM score surpassing the clinically meaningful change threshold (-4.5)
<sup>a</sup>SD, standard deviation
<sup>b</sup>AIM, Autism Impact Measure

The secondary outcome analysis did indicate a statistically significant time-by-treatment interaction for the AIM communication subdomain (Table 8). The control group showed an increase in score from baseline to the end-of-study of 3.33, indicating clinical deterioration. By contrast, the treatment group showed a decrease from baseline to the end-of-study of -5.12, consistent with clinical improvement based on MCT.

**Table 8.** Secondary Outcome Measure Analysis.

|  | Control<br>(*n* = 6) |  | Control<br>Group<br>Change<br>Score | Treatment<br>(*n* = 8) |  | Treatment<br>Group<br>Change<br>Score | Interaction F-Statistics |
| --- | --- | --- | --- | --- | --- | --- | --- |
|  | Pre-Mean<br>(SD<sup>a</sup>) | Post-Mean<br>(SD<sup>a</sup>) |  | Pre-Mean<br>(SD<sup>a</sup>) | Post-Mean<br>(SD<sup>a</sup>) |  |  |
| AIM<sup>b</sup> |  |  |  |  |  |  |  |
| Social Reciprocity | 26.00<br>(6.32) | 26.00<br>(6.87) | 0 | 23.00<br>(5.90) | 21.25<br>(5.95) | **-1.75** | 0.19 (p=0.67) |
| Communication | 35.17<br>(11.09) | 38.50<br>(8.73) | 3.33<sup>^</sup> | 30.00<br>(7.13) | 24.88<br>(6.24) | **-5.12<sup>†</sup>** | **6.75 (p=0.02)\*** |
| Peer Interaction | 25.00<br>(5.55) | 21.50<br>(8.43) | **-3.5** | 18.13<br>(6.71) | 14.13<br>(3.87) | **-4** | 0.02 (p=0.90) |
| Repetitive<br>Behavior | 38.17<br>(14.77) | 39.00<br>(15.74) | 0.83<sup>^</sup> | 38.00<br>(11.64) | 35.25<br>(12.56) | **-2.75** | 0.50 (p=0.51) |
| Atypical Behavior | 32.00<br>(11.22) | 29.67<br>(12.93) | **-2.33** | 27.13<br>(10.40) | 22.25<br>(6.04) | **-4.88** | 0.37 (p=0.55) |

\*Indicates a significant Group×Time (pre-post) interaction test statistic at $p<0.05$
<sup>†</sup>Indicates a significant estimated marginal means for post-hoc pairwise comparison test of within-group change at $p<0.05$
<sup>^</sup>Indicates a clinical deterioration (domain-dependent per Silkey et al.)<sup>47</sup>
**Bold** indicates an average decrease in AIM score surpassing the clinically meaningful change threshold (domain-dependent per Silkey et al.)<sup>47</sup>
<sup>a</sup>SD, standard deviation
<sup>b</sup>ABA, Applied Behavior Analysis

Pairwise estimated marginal mean *post hoc* tests were calculated to evaluate the significance of within-group change. Improvement in AIM communication scores for the treatment group was found to be significant (p=0.02). In addition, the treatment group showed numerical trends toward improvement (based on MCT) in the AIM scores for social reciprocity (-1.75), peer interaction (-4), repetitive behavior (-2.75), and atypical behavior (-4.88), although these did not meet the threshold for statistical significance. The control group showed trends toward improvement in peer interaction (-3.5) and atypical behaviors (-2.33). There was no trend toward change with respect to MCT in repetitive behavior (0.83) or social reciprocity (0) for the control group.

*Post hoc* between-group analyses indicated that scores on the AIM communication subdomain at end-of-study were significantly different across intervention groups (Table 9), with lower scores in the Floreo BSC treatment group compared to the control group (p=0.014). It is important to note that there were no significant group differences in these scores at baseline.

**Table 9.** End-of-study Descriptive Statistics and Group Differences.

|  | **Control**<br>(*n* = 6) | **Treatment**<br>(*n* = 8) | ***p*-Value** |
| --- | --- | --- | --- |
|  | Mean (SD<sup>a</sup>)<br>[min-max] | Mean (SD<sup>a</sup>)<br>[min-max] |  |
| AIM<sup>b</sup> |  |  |  |
| Composite | 217.83 (57.67) [133.00-276.00] | 167.88 (35.99) [137.00-252.00] | 0.2 |
| Social | 26.00 (6.87)<br>[17.00-33.00] | 21.25 (5.95)<br>[14.00-31.00] | 0.3 |
| Communication* | 38.50 (8.73)<br>[26.00-49.00] | 24.88 (6.24)<br>[17.00-36.00] | 0.014* |
| Peer Interaction | 21.50 (8.43)<br>[11.00-31.00] | 14.13 (3.87)<br>[8.00-19.00] | 0.2 |
| Repetitive Behavior | 39.00 (15.74)<br>[21.00-58.00] | 35.25 (12.56)<br>[25.00-64.00] | >0.9 |
| Atypical Behavior | 29.67 (12.93)<br>[13.00-47.00] | 22.25 (6.04)<br>[15.00-31.00] | 0.3 |

\*Indicates a significant test statistic at $p < 0.05$
<sup>a</sup>SD, standard deviation
<sup>b</sup>ABA, Applied Behavior Analysis

## Discussion

### Principal Findings

The primary goal of the present study was to examine the feasibility of using Floreo’s immersive VR program, Floreo BSC, with autistic preschool and school-aged children in a naturalistic treatment setting. Very few mild adverse events were reported, most without any clear relation to VR, and there were no serious adverse events. These findings add to the growing body of evidence supporting the safety and feasibility of immersive VR technology as a component of therapy for children with ASD.^39,40^

We also examined the efficacy of Floreo BSC for ameliorating core symptoms of autism in preschool and school-aged children, using the AIM as an outcome measure. Overall, the results showed that autistic children treated with Floreo’s immersive VR BSC program improved clinically in the AIM composite score, with a decrease of 25 points, compared to control participants who received the non-instructional VR exposure and decreased only 0.84 points on average. While numerically striking, this difference did not achieve statistical significance, likely due to the small number of participants in this feasibility study.

Despite the relatively small number of participants, we were able to demonstrate both clinically meaningful and statistically significant changes related to treatment in the AIM communication subdomain, one of the skill areas most directly targeted by the BSC intervention. Children who used Floreo BSC improved by an amount surpassing previously established meaningful change thresholds, while those exposed to a VR control appeared to show deterioration in communication scores.^47^

Children in the Floreo BSC group also showed clinical improvement in the other four subdomains of the AIM that surpassed MCT thresholds. On the other hand, children in the control group showed smaller improvements in peer interaction and repetitive behavior, and no clinically meaningful change in social reciprocity or atypical behavior. These trends suggest that Floreo BSC may be beneficial for improving core autism symptoms beyond communication.

The AIM is one of the only clinical assessment tools designed to assess both the frequency and functional impact of symptoms in autism.^46^ To the best of our knowledge, this is the first study to use the AIM to assess treatment outcomes after a VR intervention. Compared to prior studies using the AIM to measure changes in symptom severity, this study suggests larger clinically meaningful changes within a shorter period of treatment. For example, Silkey et al.^47^ estimated that a change in communication score of -0.89 over 12 months represented meaningful clinical improvement. Participants in the Floreo BSC group in this study showed a statistically significant change of -5.12 over a period of only 3 months of treatment. Similar trends toward improvement were observed for the AIM composite and other subdomain scores in the Floreo BSC treatment group.

It is tempting to interpret these results as suggesting that VR intervention may contribute to more rapid improvement in core autism symptoms than what is typically observed in the community. Of course, our ability to generalize these findings is limited by variability inherent in the small sample size. Moreover, while the AIM was designed to be sensitive to treatment effects, it is unclear whether the MCT values calculated by Silkey et al.^47^ are reliable over shorter treatment periods.

Floreo’s VR intervention is uniquely designed for paired interactions, enabling clinicians, teachers, or family members to set up lessons, monitor progress, modify the interactive environment, and provide real-time guidance, rewards, and feedback. This flexibility allows for sessions to be conducted either in a clinical setting or remotely at home, making it both cost-efficient and accessible to a large number of families of children with autism. Here, Floreo BSC was employed in a clinical setting without obvious difficulties in implementation, adherence, or tolerability.

### Comparison with Prior Work

Overall, our findings support prior studies showing that immersive VR interventions improved social skills in children with autism.^7,40,50-52^ Zhao et al.^50^ reported that children with ASD who utilized a rehabilitation therapy-based VR intervention over 3 months had improved facial recognition, happy tone of voice, and body language in response to virtual characters compared to those in the control group, as measured using the Psychoeducational Profile, 3rd Edition (PEP-3). Another study showed improved social skills scores on the PEP-3 in autistic children after one session of using an immersive, four-sided Cave Automatic Virtual Environment program.^51^ Ip et al.^52^ reported that children with autism showed significant improvement in their social interactions, adaptation skills, and emotion regulation, as measured by various assessment scales, after using the Cave Automatic Virtual Environment intervention for 14 weeks. Additionally, Frolli et al.^7^ demonstrated autistic children who used an immersive emotional literacy VR intervention had an enhanced ability to recognize different types of emotions.

### Limitations

As we have already noted, conclusions about the efficacy of the Floreo BSC intervention are limited by the small number of subjects included in this study, which was primarily designed to assess feasibility. We have also discussed potential issues with using the AIM as a short-term measurement tool; it may be more appropriate in future studies to assess responder rates rather than aggregate scores. Future studies may also be strengthened by examining additional secondary outcome measures, including measures that are already widely employed clinically, such as the Vineland Adaptive Behavior Scales.

More broadly, there is the potential for participant self-selection bias in studies of VR-based interventions. In other words, participants who had a pre-existing positive view of VR and similar technologies may have been more inclined to enroll in the study. Clinicians and educators should be mindful of individual preferences and differences when participating in VR.^53^

## Conclusions

Overall, this study suggests that Floreo’s immersive VR Building Social Connections Module is safe and well-tolerated. The BSC curriculum may lead to both clinical and statistically significant improvements in social communication skills in school-aged children with autism, compared to children who received a VR exposure control, as evidenced by improvements in AIM communication scores. This study also observed overall clinical improvement for the treatment group in the AIM composite score as well as in the social reciprocity, peer interaction, and repetitive and atypical behavior subdomains, suggesting that Floreo’s immersive VR intervention may be broadly beneficial in ameliorating symptoms associated with autism.

## Data Availability

All data produced in the present study are available upon reasonable request to the authors

## Acknowledgments

The authors wish to thank the following:

1. All participants and families who participated in this study.
2. Study teams who ran the study.

## Authorship Confirmation/Contribution Statement

**Kevin Shapiro**: Conceptualization; methodology; writing - original draft, review and editing; supervision.

**Vijay Ravindran**: Conceptualization; methodology; validation; software; writing - original draft, review and editing; funding acquisition; supervision.

**Gregory Downing**: Conceptualization; methodology; resources; writing - original draft, review and editing; funding acquisition; supervision.

**Shirley Mak-Parisi**: Data curation; formal analysis; project administration; visualization; writing - original draft, review and editing, supervision.

**Georgia Barbayannis**: Visualization; validation; writing - review and editing.

**Sinan Turnacioglu**: Conceptualization; methodology; writing - original draft, review and editing; supervision.

## Clinical Trials Registration Information

NCT05791071

## Conflicts of Interest

Vijay Ravindran is the Chief Executive Officer and cofounder of Floreo, Inc.

Shirley Mak-Parisi is an employee of Floreo, Inc.

Greg Downing and Sinan Turnacioglu are consultants of Floreo, Inc.

## Funding Statement

This is research sponsored by Floreo, Inc.

## Abbreviations

ABA: Applied behavior analysis
AIM: Autism Impact Measure
ASD: Autism spectrum disorder
BSC: Building Social Connections
CARS-2: Childhood Autism Rating Scale, 2nd Edition
CI: clinical improvement
ITT: intent-to-treat
KBIT: Kaufman Brief Intelligence Test, 2nd Edition
MCT: Meaningful change threshold
MT: music therapy
OT: occupational therapy
PEP-3: Psychoeducational Profile, 3rd Edition
PT: physical therapy
Vineland-3: Vineland Scales of Adaptive Behavior, 3rd Edition
VR: Virtual reality

## Notes

### Clinical Trial

NCT05791071

### Author Declarations

WCG, Central Internal Review Board of Cortica gave ethical approval for this work.

## References

1. American Psychiatric Association. Diagnostic and Statistical Manual of Mental Disorders, DSM-5. Fifth Edition. Washington, DC: American Psychiatric Association; 2013.

2. van Heijst BF, Geurts HM. Quality of life in autism across the lifespan: a meta-analysis. Autism. 2015;19(2):158–67. doi: 10.1177/1362361313517053. PMID: 24443331.

3. Warreman EB, Lloyd SE, Nooteboom LA, Leenen PJM, Terry MB, Hoek HW, van Rossum EFC, Vermeiren RRJM, Ester WA. Psychological, behavioural, and physical aspects of caregiver strain in autism-caregivers: a cohort study. EClinicalMedicine. 2023;64:102211. doi: 10.1016/j.eclinm.2023.102211. PMID: 37767192; PMCID: PMC10520302.

4. Pecor K, Barbayannis G, Yang M, Johnson J, Materasso S, Borda M, Garcia D, Garla V, Ming X. Quality of life changes during the COVID-19 pandemic for caregivers of children with ADHD and/or ASD. International Journal of Environmental Research and Public Health 2021;18(7):3667. doi: 10.3390/ijerph18073667

5. Maenner MJ, Warren Z, Williams A, Amoakohene E, Bakian A, Bilder D, Durkin M, Fitzgerald R, Furnier S, Hughes M, Ladd-Acosta C, McArthur D, Pas E, Salinas A, Vehorn A, Williams S, Esler A, Grzybowski A, Hall-Lande J, Nguyen R. Prevalence and characteristics of autism spectrum disorder among children aged 8 years—autism and developmental disabilities monitoring network, 11 sites, United States. MMWR. Surveillance Summaries 2023;72(2):1–14. doi: 10.15585/mmwr.ss7202a1

6. Kenny L, Hattersley C, Molins B, Buckley C, Povey C, Pellicano E. Which terms should be used to describe autism? perspectives from the UK autism community. Autism. 2016;20(4):442–462. doi: 10.1177/1362361315588200

7. Frolli A, Savarese G, Di Carmine F, Bosco A, Saviano E, Rega A, Carotenuto M, Ricci MC. Children on the autism spectrum and the use of virtual reality for supporting social skills. Children. 2022;9(2):181. doi: 10.3390/children9020181

8. Schreibman L, Dawson G, Stahmer AC, Landa R, Rogers SJ, McGee GG, Kasari C, Ingersoll B, Kaiser AP, Bruinsma Y, McNerney E, Wetherby A, Halladay A. Naturalistic developmental behavioral interventions: empirically validated treatments for autism spectrum disorder. Journal of Autism and Developmental Disorders. 2015;45(8):2411–2428. doi: 10.1007/s10803-015-2407-8

9. Rogers SJ, Vismara LA. Evidence-based comprehensive treatments for early autism. Journal of Clinical Child & Adolescent Psychology. 2008;37(1):8–38. doi: 10.1080/15374410701817808

10. Tiede G, Walton KM. Meta-analysis of naturalistic developmental behavioral interventions for young children with autism spectrum disorder. Autism: The International Journal of Research and Practice. 2019;23(8):2080–2095. doi: 10.1177/1362361319836371

11. Cidav Z, Munson J, Estes A, Dawson G, Rogers S, Mandell D. Cost offset associated with early start Denver model for children with autism. Journal of the American Academy of Child & Adolescent Psychiatry. 2017;56(9):777–783. doi: 10.1016/j.jaac.2017.06.007

12. Janvier YM, Harris JF, Coffield CN, Louis B, Xie M, Cidav Z, Mandell DS. Screening for autism spectrum disorder in underserved communities: early childcare providers as reporters. Autism. 2016;20(3):364–373. doi: 10.1177/1362361315585055

13. Malik-Soni N, Shaker A, Luck H, Mullin AE, Wiley RE, Lewis MES, Fuentes J, Frazier TW. Tackling healthcare access barriers for individuals with autism from diagnosis to adulthood. Pediatric Research. 2022;91(5):1028–1035. doi: 10.1038/s41390-021-01465-y

14. Liu BM, Paskov K, Kent J, McNealis M, Sutaria S, Dods O, Harjadi C, Stockham N, Ostrovsky A, Wall DP. Racial and ethnic disparities in geographic access to autism resources across the US. JAMA Network Open. 2023;6(1):e2251182. doi: 10.1001/jamanetworkopen.2022.51182

15. Aylward BS, Gal-Szabo DE, Taraman S. Racial, ethnic, and sociodemographic disparities in diagnosis of children with autism spectrum disorder. Journal of Developmental & Behavioral Pediatrics. 2021;42(8):682–689. doi: 10.1097/dbp.0000000000000996

16. Pellecchia M, Connell JE, Kerns CM, Xie M, Marcus SC, Mandell DS. Child characteristics associated with outcome for children with autism in a school-based behavioral intervention. Autism, 2016;20(3):321–329. doi: 10.1177/1362361315577518

17. Nahmias AS, Kase C, Mandell DS. Comparing cognitive outcomes among children with autism spectrum disorders receiving community-based early intervention in one of three placements. Autism. 2014;18(3):311–320. doi: 10.1177/1362361312467865

18. Howlin P, Savage S, Moss P, Tempier A, Rutter M. Cognitive and language skills in adults with autism: a 40-year follow-up. Journal of Child Psychology and Psychiatry. 2014;55(1):49–58. doi: 10.1111/jcpp.12115

19. Ames JL, Morgan EH, Giwa Onaiwu M, Qian Y, Massolo ML, Croen LA. Racial/ethnic differences in psychiatric and medical diagnoses among autistic adults. Autism in Adulthood. 2022;4(4):290–305. doi: 10.1089/aut.2021.0083

20. Eaves LC, Ho HH. Young adult outcome of autism spectrum disorders. Journal of Autism and Developmental Disorders. 2008;38(4):739–747. doi: 10.1007/s10803-007-0441-x

21. Cadman T, Eklund H, Howley D, Hayward H, Clarke H, Findon J, Xenitidis K, Murphy D, Asherson P, Glaser K. Caregiver burden as people with autism spectrum disorder and attention-deficit/hyperactivity disorder transition into adolescence and adulthood in the United Kingdom. Journal of the American Academy of Child & Adolescent Psychiatry. 2012;51(9):879–888. doi: 10.1016/j.jaac.2012.06.017

22. Orsmond GI, Shattuck PT, Cooper BP, Sterzing PR, Anderson KA. Social participation among young adults with an autism spectrum disorder. Journal of Autism and Developmental Disorders. 2013;43(11):2710–2719. doi: 10.1007/s10803-013-1833-8

23. Garcha J, Smith AP. Associations between autistic and ADHD traits and the well-being and mental health of university students. Healthcare Multidisciplinary Digital Publishing Institute. 2023;12(1):14–28. doi: 10.3390/healthcare12010014

24. Bougeard C, Picarel-Blanchot F, Schmid R, Campbell R, Buitelaar J. Prevalence of autism spectrum disorder and co-morbidities in children and adolescents: a systematic literature review. Focus/Focus (American Psychiatric Publishing). 2024;22(2):212–228. doi: 10.1176/appi.focus.24022005

25. Roux AM, Shattuck PT, Rast JE, Rava JA, Edwards AD, Wei X, McCracken M, Yu JW. Characteristics of two-year college students on the autism spectrum and their support services experiences. Autism Research and Treatment. 2015;2015:391693. doi: 10.1155/2015/391693

26. Wallace GL, Budgett J, Charlton RA. Aging and autism spectrum disorder: evidence from the broad autism phenotype. Autism Research. 2016;9(12):1294–1303. doi: 10.1002/aur.1620

27. Pugliese CE, Anthony L, Strang JF, Dudley K, Wallace GL, Kenworthy L. Increasing adaptive behavior skill deficits from childhood to adolescence in autism spectrum disorder: role of executive function. Journal of Autism and Developmental Disorders. 2015;45(6):1579–1587. doi: 10.1007/s10803-014-2309-1

28. Damiano CR, Mazefsky CA, White SW, Dichter GS. Future directions for research in autism spectrum disorders. Journal of Clinical Child & Adolescent Psychology. 2014;43(5):828–843. doi: 10.1080/15374416.2014.945214

29. Roche L, Adams D, Clark M. Research priorities of the autism community: a systematic review of key stakeholder perspectives. Autism. 2020;25(2):336–348. doi: 10.1177/1362361320967790

30. Parsons S, Cobb S. State-of-the-art of virtual reality technologies for children on the autism spectrum. European Journal of Special Needs Education. 2011;26(3):355–366. doi: 10.1080/08856257.2011.593831

31. Ramsey KA, Essoe JK-Y, Boyle N, Patrick AK, McGuire JF. Immersive virtual reality exposures for the treatment of childhood anxiety. Child Psychiatry & Human Development. 2023;1–12. doi: 10.1007/s10578-023-01628-4

32. Shin B, Oh J, Kim B-H, Kim HE, Kim H, Kim S-J, Kim J-J. Effectiveness of self-guided virtual reality-based cognitive behavioral therapy for panic disorder. JMIR Mental Health. 2021;8(11):e30590. doi: 10.2196/30590

33. Donnelly MR, Reinberg R, Ito KL, Saldana D, Neureither M, Schmiesing A, Jahng E, Liew S-L. Virtual reality for the treatment of anxiety disorders: a scoping review. The American Journal of Occupational Therapy. 2021;75(6):7506205040. doi: 10.5014/ajot.2021.046169

34. Lahiri U, Bekele E, Dohrmann E, Warren Z, Sarkar N. A physiologically informed virtual reality based social communication system for individuals with autism. Journal of Autism and Developmental Disorders. 2015;45(4):919–931. doi: 10.1007/s10803-014-2240-5

35. Didehbani N, Allen T, Kandalaft M, Krawczyk D, Chapman S. Virtual reality social cognition training for children with high functioning autism. Computers in Human Behavior. 2016;62:703–711. doi: 10.1016/j.chb.2016.04.033

36. Josman N, Ben-Chaim HM, Friedrich S, Weiss PL. Effectiveness of virtual reality for teaching street-crossing skills to children and adolescents with autism. International Journal on Disability and Human Development. 2008;7(1):49–56. doi: 10.1515/ijdhd.2008.7.1.49

37. Odom SL, Thompson JL, Hedges S, Boyd BA, Dykstra JR, Duda MA, Szidon KL, Smith LE, Bord A. Technology-aided interventions and instruction for adolescents with autism spectrum disorder. Journal of Autism and Developmental Disorders. 2015;45(12):3805–3819. doi: 10.1007/s10803-014-2320-6

38. Zhang M, Ding H, Naumceska M, Zhang Y. Virtual reality technology as an educational and intervention tool for children with autism spectrum disorder: current perspectives and future directions. Behavioral Sciences. 2022;12(5):138. doi: 10.3390/bs12050138

39. Ravindran V, Osgood M, Sazawal V, Solorzano R, Turnacioglu S. Virtual reality support for joint attention using the Floreo Joint Attention Module: usability and feasibility pilot study. JMIR Pediatrics and Parenting. 2019;2(2):e14429. doi: 10.2196/14429

40. Mittal P, Bhadania M, Tondak N, Ajmera P, Yadav S, Kukreti A, Kalra S, Ajmera P. Effect of immersive virtual reality-based training on cognitive, social, and emotional skills in children and adolescents with autism spectrum disorder: a meta-analysis of randomized controlled trials. Research in Developmental Disabilities Elsevier BV. 2024;151:104771. doi: 10.1016/j.ridd.2024.104771

41. Wainer AL, Ingersoll BR. The use of innovative computer technology for teaching social communication to individuals with autism spectrum disorders. Research in Autism Spectrum Disorders. 2011;5(1):96–107. doi: 10.1016/j.rasd.2010.08.002

42. Mesa-Gresa P, Gil-Gómez H, Lozano-Quilis J-A, Gil-Gómez J-A. Effectiveness of virtual reality for children and adolescents with autism spectrum disorder: an evidence-based systematic review. Sensors. 2018;18(8):2486. doi: 10.3390/s18082486

43. Lorenzo G, Lledó A, Pomares J, Roig R. Design and application of an immersive virtual reality system to enhance emotional skills for children with autism spectrum disorders. Computers & Education. 2016;98:192–205. doi: 10.1016/j.compedu.2016.03.018

44. Kanne SM, Mazurek MO, Sikora D, Bellando J, Branum-Martin L, Handen B, Katz T, Freedman B, Powell MP, Warren Z. The autism impact measure (AIM): initial development of a new tool for treatment outcome measurement. Journal of Autism and Developmental Disorders. 2014;44(1):168–179. doi: 10.1007/s10803-013-1862-3

45. Houghton R, Monz B, Law K, Loss G, Le Scouiller S, de Vries F, Willgoss T. Psychometric validation of the autism impact measure (AIM). Journal of Autism and Developmental Disorders. 2019;49(6):2559–2570. doi: 10.1007/s10803-019-04011-2

46. Mazurek MO, Carlson C, Baker-Ericzén M, Butter E, Norris M, Barr C, Kanne S. The autism impact measure (AIM): examination of sensitivity to change. Autism Research: Official Journal of the International Society for Autism Research. 2020;13(11):1867–1879. doi: 10.1002/aur.2397

47. Silkey M, Durán-Pacheco G, Johnson M, Liu C, Clinch S, Law K, Loss G. The autism impact measure (AIM): meaningful change thresholds and core symptom changes over one year from an online survey in the U.S. Journal of Autism and Developmental Disorders. 2023;53(9):3422–3434. doi: 10.1007/s10803-022-05635-7

48. Sparrow SS, Cicchetti DV, Saulnier CA. Vineland adaptive behavior scales. 3rd ed. San Antonio: TX, Pearson; 2016. ISBN:9781488694127

49. Chatham CH, Taylor KI, Charman T, Liogier D’ardhuy X, Eule E, Fedele A, Hardan AY, Loth E, Murtagh L, del Valle Rubido M, San Jose Caceres A, Sevigny J, Sikich L, Snyder L, Tillmann JE, Ventola PE, Walton-Bowen KL, Wang PP, Willgoss T, Bolognani F. Adaptive behavior in autism: minimal clinically important differences on the Vineland-II. Autism Research. 2018;11(2):270–283. doi: 10.1002/aur.1874

50. Zhao J, Zhang X, Lu Y, Wu X, Zhou F, Yang S, Wang L, Wu X, Fei F. Virtual reality technology enhances the cognitive and social communication of children with autism spectrum disorder. Frontiers in Public Health. 2022;10:1029392. doi: 10.3389/fpubh.2022.1029392

51. Yuan SNV, Ip HHS. Using virtual reality to train emotional and social skills in children with autism spectrum disorder. London Journal of Primary Care. 2018;10(4):110–112. doi: 10.1080/17571472.2018.1483000

52. Ip HHS, Wong SWL, Chan DFY, Byrne J, Li C, Yuan VSN, Lau KSY, Wong JYW. Enhance emotional and social adaptation skills for children with autism spectrum disorder: a virtual reality enabled approach. Computers & Education. 2018;117:1–15. doi: 10.1016/j.compedu.2017.09.010

53. McCleery JP, Zitter A, Solórzano R, Turnacioglu S, Miller JS, Ravindran V, Parish Morris J. Safety and feasibility of an immersive virtual reality intervention program for teaching police interaction skills to adolescents and adults with autism. Autism Research. 2020;13(8):1418–1424. doi: 10.1002/aur.2352

